# Plasma Phosphorylated Tau 217 Varies With Kidney Function Even Without Chronic Kidney Disease

**DOI:** 10.64898/2026.10.18.26365853

**Authors:** Matthew JY Kang, Tom Borchard, Michelle M. Mielke, Henrik Zetterberg, Malcolm Hopwood, Phillip B Mitchell, Dhamidhu Eratne, Dennis Velakoulis

## Abstract

**Importance:** Plasma phosphorylated tau 217 (p-tau217) is entering clinical use with fixed diagnostic threshold that do not account for kidney function. Chronic kidney disease (defined as estimated glomerular filtration rate (eGFR) below 60 mL/min/1.73 m^2^), raises plasma p-tau217 concentrations and can produce false positive results. It remains unclear whether milder reductions in kidney function, above the CKD threshold, have the same effect.

**Objective:** To determine whether plasma p-tau217 is elevated with mildly reduced kidney function, above the chronic kidney disease threshold, in adults without neurological disease.

**Design:** Cross-sectional analysis of a cohort enrolled between 2023 and 2025 and biomarker analysis blinded to clinical data.

**Setting:** Two private psychiatric inpatient facilities in Australia.

**Participants:** Adults admitted with psychiatric disorders and free of neurological disease.

**Exposures:** eGFR calculated from serum creatinine using the CKD-EPI 2021 equation.

**Main Outcomes and Measures:** Plasma p-tau217, neurofilament light chain (NfL), and glial fibrillary acidic protein (GFAP), measured on the Simoa platform. Multivariable linear regression compared eGFR groups (< 60, 60 to 90, and ≥ 90 mL/min/1.73 m^2^), adjusted for age, BMI, and sex.

**Results:** Among 180 participants (median age, 50.5 years [IQR, 38.0-61.5]; 94 [52%] female), eGFR was ≥ 90 mL/min/1.73 m^2^ in 106 (59%), 60 to 90 in 61 (34%), and < 60 in 13 (7.2%). Compared with an eGFR ≥ 90, plasma p-tau217 was 36% higher in both the 60 to < 90 group (β = 0.42 [95% CI: 0.11, 0.74]; P = .008) and 116% higher in the < 60 group (β = 1.05 [95% CI: 0.43, 1.72]; P = .002). Plasma NfL and GFAP were elevated in the < 60 group but differences in the 60 to < 90 group did not reach significance. Modelled continuously, plasma p-tau217 was 9.5% lower per 10 mL/min/1.73 m^2^ higher eGFR (95% CI: 3.3%, 15.4%).

**Conclusions and Relevance:** Plasma p-tau217 was elevated in adults with mildly reduced kidney function, above the CKD threshold, in a cohort free of neurological disease. Kidney function across its full range should be incorporated into plasma p-tau217 reference ranges and clinical interpretation to reduce false positive and false negative results.

**KEY POINTS:** *Question:* Is plasma p-tau217 elevated at mildly reduced kidney function, above the chronic kidney disease (CKD) threshold?

*Findings:* In this cross-sectional study of 180 inpatients free of neurological disease, plasma p-tau217 was 36% higher in participants with an estimated glomerular filtration rate (eGFR) between 60 and 90 mL/min/1.73 m^2^, and 116% higher in those below 60, compared with those at 90 or above.

*Meaning:* Given that plasma p-tau217 varies with kidney function across its range, not only in CKD, eGFR should be incorporated into reference ranges and the interpretation of results to reduce false positive and false negative findings.

## INTRODUCTION

Plasma phosphorylated tau 217 (p-tau217) has entered clinical use internationally.^1,2^ Interpretation relies on fixed reference intervals, which assume concentrations reflect brain pathology rather than peripheral physiology. That assumption is imperfect. Plasma p-tau217, neurofilament light chain (NfL), and glial fibrillary acidic protein (GFAP) are elevated in chronic kidney disease (CKD) and correlate inversely with filtration rate,^3–6^ thus caution is advised in interpreting blood-based biomarkers in severe CKD.^7^

CKD is defined as an estimated glomerular filtration rate (eGFR) below 60 mL/min/1.73 m^2^, and this threshold has been the focus of the existing evidence and guidelines.^7–9^ Plasma p-tau217 has been reported to be affected only below 60 mL/min/1.73 m^2^,^6,10^ and the largest study to date stratified participants at 60 mL/min/1.73 m^2^ and subdivided only downwards, leaving the mildly reduced range (i.e. 60-90 mL/min/1.73 m^2^) unexamined.^11^ Whether plasma p-tau217 is elevated above the defined CKD threshold (i.e., >60 mL/min/1.73 m^2^), a range that would not prompt clinical concern, has not been directly tested.

We compared plasma p-tau217, NfL, and GFAP across groups of kidney function, with particular attention to the mildly reduced range above the CKD threshold, in adults free of neurological disease.

## METHODS

This cross-sectional study analysed participants enrolled between 2023 and 2025 in a prospective cohort at private psychiatric inpatient facilities in Australia as part of The Markers in Neuropsychiatric Disorders Study (MiND Study).^12^ Eligible participants were adult inpatients who did not have comorbid neurological disease. eGFR used the CKD-EPI 2021 creatinine equation.^13^ The MiND Study was approved by the local ethics committees (MH2020.142). Participants provided written consent and reporting follows STROBE.

### Blood sample collection and biomarker analysis

EDTA plasma collected at admission was stored at −80 °C. Biomarkers were analysed on Quanterix Simoa HD-X (NfL and GFAP using N2PB assay; p-tau217 using in-house University of Gothenburg assay.^14^

### Statistical analysis

The primary analysis compared eGFR groups, defined as below 60, 60 to under 90, and 90 or above mL/min/1.73 m^2^ in accordance with international guidance and clinical reporting.^8^ Multivariable linear regression adjusted for age, BMI, and sex, with 90 or above as the reference. eGFR was also modelled continuously to assess whether the association was linear without categorical decreases. Confidence intervals were derived from 10,000 bootstrap resamples. Coefficients are reported as standardised β and as percentage difference from the reference group.

Eleven GFAP and three p-tau217 values were below the lower limit of quantification. All were included in the primary models and treated as missing in sensitivity analyses.

As age is an input to the CKD-EPI equation, we anticipated age and eGFR to be correlated (r = −0.67, VIF = 1.84) and conducted two additional analyses assessing age: models predicting creatinine instead of eGFR, and models restricted to participants aged 45 to 70 years.

## RESULTS

Among 180 participants (median age, 50.5 years [IQR, 38.0-61.5]; 94 [52%] female; 127 [71%] with major depressive disorder), eGFR was 90 mL/min/1.73 m^2^ or above in 106 (59%), 60 to under 90 in 61 (34%), and below 60 in 13 (7.2%). Age differed across eGFR groups (median, 40.0, 59.0, and 67.0 years; P < .001), whereas other characteristics including cognition (MOCA) did not (Table 1). Unadjusted plasma p-tau217 levels increased with decreasing eGFR group (Figure 1).

**Table 1.** Participant characteristics and unadjusted plasma biomarker concentrations by eGFR category (CKD-EPI 2021).

| Characteristic | Overall<br>N = 180 | < 60<br>n = 13 | 60–90<br>n = 61 | ≥ 90<br>n = 106 | P<br>value |
| --- | --- | --- | --- | --- | --- |
| Age at blood draw (years) | 50.5 (38.0, 61.5) | 67.0 (59.0, 76.0) | 59.0 (52.0, 71.0) | 40.0 (32.0, 51.0) | <.001 |
| Sex, female | 94 (52%) | 8 (62%) | 30 (49%) | 56 (53%) | .70 |
| BMI (kg/m<sup>2</sup>) | 28.7 (24.4, 32.8) | 27.7 (25.6, 34.8) | 29.1 (25.1, 32.3) | 28.4 (24.2, 32.9) | >.90 |
| Primary diagnosis |  |  |  |  | .70 |
| Major depressive disorder | 127 (71%) | 8 (62%) | 43 (70%) | 76 (72%) |  |
| Bipolar disorder | 53 (29%) | 5 (38%) | 18 (30%) | 30 (28%) |  |
| MoCA | 27.0 (24.0, 28.0) | 27.0 (24.0, 29.0) | 26.0 (24.0, 28.0) | 27.0 (25.0, 28.0) | .70 |
| Plasma p-tau217 (pg/mL) | 1.39 (0.95, 1.87) | 2.65 (1.67, 2.96) | 1.48 (1.12, 2.39) | 1.17 (0.90, 1.61) | <.001 |
| Plasma NfL (pg/mL) | 7.88 (5.92, 11.61) | 21.81 (17.79, 23.87) | 10.58 (7.15, 16.39) | 6.50 (4.84, 9.27) | <.001 |
| Plasma GFAP (pg/mL) | 86.10 (56.90, 126.50) | 159.00 (122.00, 196.00) | 119.00 (86.30, 166.00) | 66.30 (51.40, 97.40) | <.001 |

Values are median (Q1, Q3) or No. (%). P values from Kruskal-Wallis rank sum test, Pearson chi-squared test, or Fisher exact test as appropriate. Montreal Cognitive Assessment (MoCA) missing for 7, ACE-III for 22, and APOE ε4 dose for 61 participants. eGFR units are mL/min/1.73 m<sup>2</sup>.
BMI = body-mass index, GFAP = glial fibrillary acidic protein, MoCA =Montreal Cognitive Assessment, NfL = neurofilament light chain, p-Tau217 = phosphorylated-Tau 217

**Figure 1.**
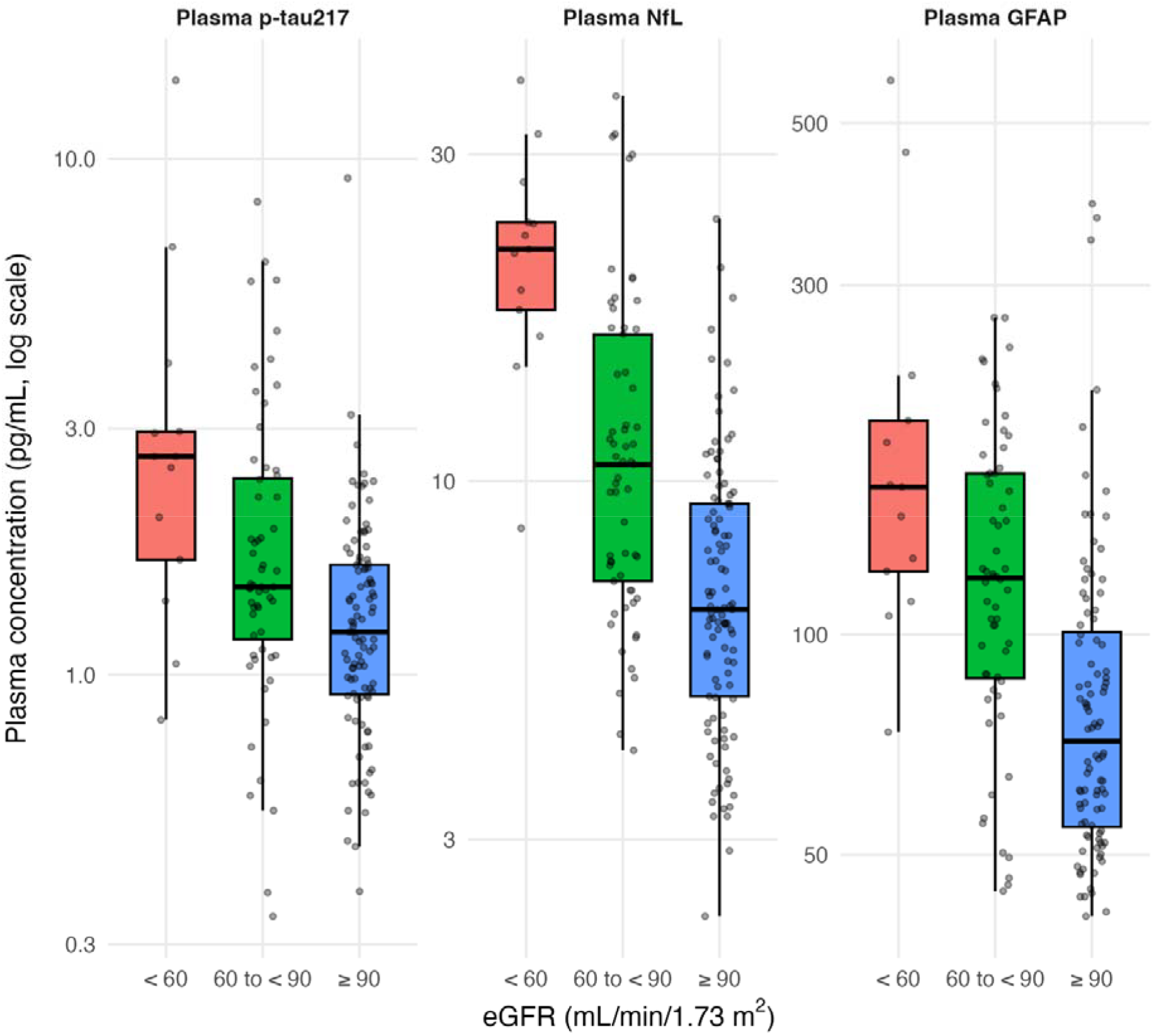
Plasma p-tau217, NfL, and GFAP concentrations by eGFR category (CKD-EPI 2021). Boxes show the median and interquartile range, whiskers extend to 1.5 times the interquartile range, and jittered points represent individual participants (eGFR < 60, n = 13; 60 to < 90, n = 61; ≥ 90, n = 106). Concentrations are plotted on a logarithmic scale. Values below the lower limit of quantification (11 plasma GFAP and three plasma p-tau217) are not displayed, and the corresponding panels therefore contain fewer points than the group sizes shown.

In the adjusted models, compared with participants with an eGFR of 90 or above, plasma p-tau217 was higher in those with an eGFR of 60 to <90 (β = 0.42 [95% CI: 0.11, 0.74]; P = .008; 36% higher [95% CI: 5%, 75%]) and elevated even more in those with an eGFR below 60 (β = 1.05 [95% CI: 0.43, 1.72]; P = .002; 116% higher [95% CI: 38%, 238%]). Within this model, in which age contributes to eGFR, age was not independently associated with plasma p-tau217 (β = 0.10 [95% CI: −0.09, 0.27]; P = .29). Substituting creatinine for eGFR, higher concentrations were associated with higher plasma p-tau217 (β = 0.30 [95% CI: 0.14, 0.45]; P < .001), exceeding the association with age in the same model (β = 0.19 [95% CI: 0.04, 0.33]; P = .011). Among participants aged 45 to 70 years, plasma p-tau217 remained higher at for an eGFR of 60 to <90 compared to 90 and above (β = 0.62 [95% CI: 0.15, 1.01]; P = .012; n = 79).

Both plasma NfL and GFAP were elevated below 60 mL/min/1.73 m^2^ (plasma NfL: β = 1.06 [95% CI: 0.60, 1.52], P < .001; plasma GFAP: β = 0.63 [95% CI: 0.17, 1.16], P = .005). At 60 to under 90, the estimates were smaller and their intervals included the null (plasma NfL: β = 0.24 [95% CI: −0.04, 0.53]; plasma GFAP: β = 0.29 [95% CI: −0.01, 0.58]).

Modelled continuously (Figure 2), higher eGFR was associated with lower plasma p-tau217 (β = −0.28 [95% CI: −0.47, −0.09]), a 9.5% lower concentration per 10 mL/min/1.73 m^2^ higher eGFR (95% CI: 3.3%, 15.4%). Comparable associations were seen for plasma NfL and GFAP (Figure 2).

**Figure 2.**
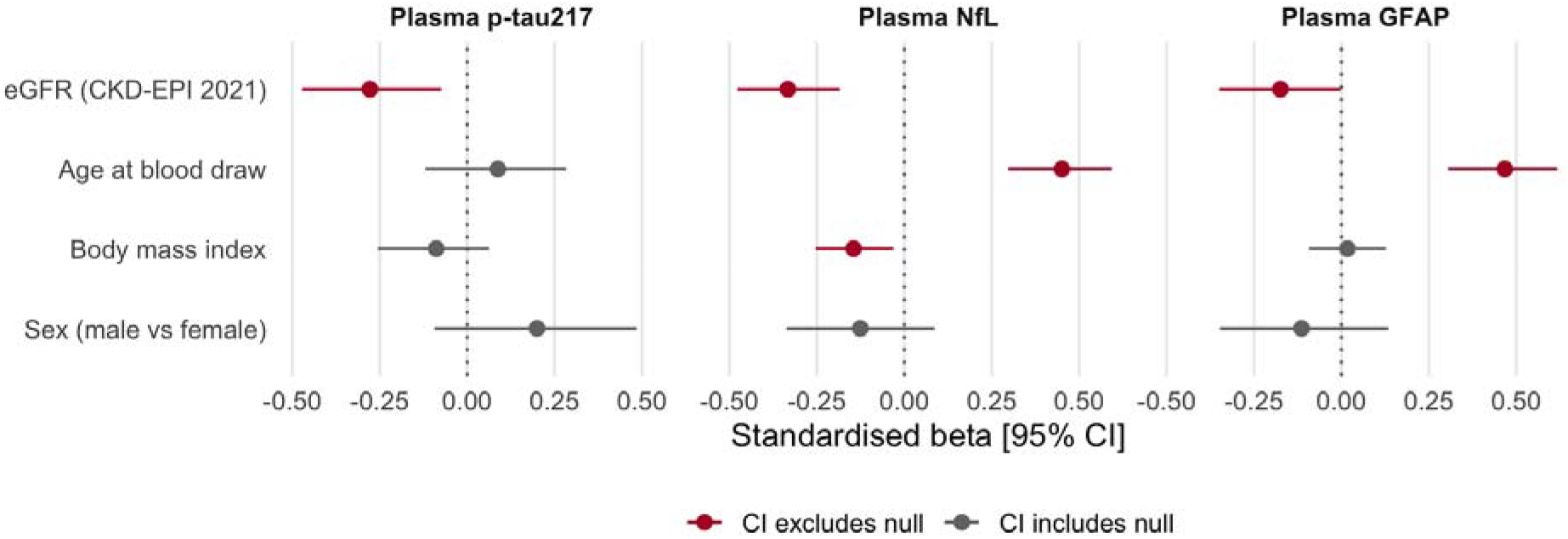
Standardised coefficients from multivariable linear models of each plasma biomarker across the full eGFR range (n = 180). Each outcome was the standardised log-transformed concentration of the biomarker named in the panel, with age at blood draw, body mass index, and eGFR (CKD-EPI 2021) entered as standardised continuous predictors and sex as a categorical predictor (reference: female). Confidence intervals are equal-tailed bootstrap intervals from 10,000 resamples. The vertical dotted line marks the null.

### Sensitivity analyses

The CKD-EPI 2009 equation gave near-identical results (r = 0.999). Treating values below the lower limit of quantification as missing did not alter the findings. The association was consistent in direction across the full eGFR range (supplemental Figure 1), and not an artefact of the eGFR cutoffs.

## DISCUSSION

In adults without neurological disease, plasma p-tau217 concentrations were significantly higher in participants with impaired renal function, including those in the mild range (eGFR between 60 to 90 mL/min/1.73 m^2^) not meeting the threshold for CKD. Existing studies of renal function and p-tau217 have focused on the typical threshold used to diagnosis CKD (eGFR < 60 mL/min/1.73 m^2^),^9,10^ and consequently, Alzheimer’s Association Clinical Practice Guideline advises caution among individuals with severe CKD only.^7^ Our findings indicate that plasma p-tau217 concentrations are also increased between 60 and 90 mL/min/1.73 m^2^ (approximately one-third higher), which could result in a falsely elevated p-tau217 concentrations.

Findings across the literature have been inconsistent. In a memory clinic cohort using the same three eGFR groups, plasma p-tau217 was affected only below 60 mL/min/1.73 m^2^.^3^ In a population-based study of 2,279 dementia-free older adults, spline models identified a rise in plasma p-tau217 below 74 mL/min/1.73 m^2^.^6^ Our results are consistent with the latter across the eGFR range, and include in a younger cohort of patients without neurodegenerative disease in which renal effects have been assumed negligible.^9,15^

Several factors may explain the discrepancy. The two cohorts used different p-tau217 assays, and the magnitude of renal effects differs substantially between platforms measured in the same participants.^10^ Comparisons in the memory clinic cohort were unadjusted, with a reference group eight years younger on average.^3^

A strength of this study is the clinical setting and the absence of neurological disease, which limits the variable contribution of neurological pathology to biomarker concentrations. Limitations include the cross-sectional design and creatinine-based estimation, which is least precise at high filtration rates. Age and eGFR are related, though the association persisted in creatinine-based models and when analysis was restricted to an age range in which the groups overlapped.

## Conclusions

In adults without neurological disease, plasma p-tau217 was elevated at mildly reduced kidney function, before the CKD threshold. Kidney function should therefore be considered whenever plasma p-tau217 is interpreted clinically, and not only when CKD is present.

## Data Availability

Data are available upon reasonable request to the authors

## Acknowledgements

The authors sincerely thank the participants of the study for their generous input for the study.

## Author Contributions

MK had full access to all the data in the study and takes responsibility for the integrity of the data and the accuracy of the data analysis.

Concept and design: MK, MM and DE

Acquisition, analysis, or interpretation of data: All authors.

Drafting of the manuscript: All authors.

Statistical analysis: MK

Critical revision of the manuscript for important intellectual content: All authors

Supervision: DV and DE

## Conflicts of Interest Disclosures

Dr. Mielke has served on scientific advisory boards and/or has consulted for Abbott, Beckman Coulter, Biogen, Eisai, Lilly, Merck, Neurogen Biomarking, Novo Nordisk, Roche, and Siemens Healthineers.

Dr. Zetterberg has served at scientific advisory boards and/or as a consultant for Abbvie, Acumen, Alamar, Alector, Alzinova, ALZpath, Amylyx, Annexon, Apellis, Artery Therapeutics, AZTherapies, Bioventix, Cognitact, Cognito Therapeutics, CogRx, Denali, Eisai, Enigma, Johnson & Johnson, LabCorp, Merck Sharp & Dohme, Merry Life, Nervgen, Neurocode, New Amsterdam, Novo Nordisk, Optoceutics, Passage Bio, Pinteon Therapeutics, Prothena, Quanterix, Red Abbey Labs, reMYND, Roche, Samumed, ScandiBio Therapeutics AB, Siemens Healthineers, Triplet Therapeutics, and Wave, has given lectures sponsored by Alzecure, BioArctic, Biogen, Cellectricon, Fujirebio, LabCorp, Lilly, Novo Nordisk, Oy Medix Biochemica AB, Roche, and WebMD, is a co-founder of Brain Biomarker Solutions in Gothenburg AB (BBS), which is a part of the GU Ventures Incubator Program, and is a shareholder of CERimmune Therapeutics (outside submitted work). The remaining authors have no conflicts of interest to declare.

## Funding/Support

This work was funded by the Ramsay Hospital Research Foundation.

MK is supported by the Research Training Program Scholarship from the Department of Psychiatry, University of Melbourne with contributions from the Australian Commonwealth Government, the Ramsay Hospital Research Foundation, and the Royal Melbourne Hospital Foundation.

HZ is a Wallenberg Scholar and a Distinguished Professor at the Swedish Research Council supported by grants from the Swedish Research Council (#2023-00356, #2022-01018 and #2019- 02397), the European Union’s Horizon Europe research and innovation programme under grant agreement No 101053962, and Swedish State Support for Clinical Research (#ALFGBG-71320). PBM was supported by an NHMRC Investigator Grant (1177991)

## Data Sharing Statement

De-identified data may be made available to researchers on reasonable request.

## Notes

### Author Declarations

The MiND Study was approved by The Royal Melbourne Hospital Human Research Ethics Committee (MH2020.142).

