## Supplemental Data 1 for "Plasma Phosphorylated Tau 217 Varies With Kidney Function Even Without Chronic Kidney Disease"

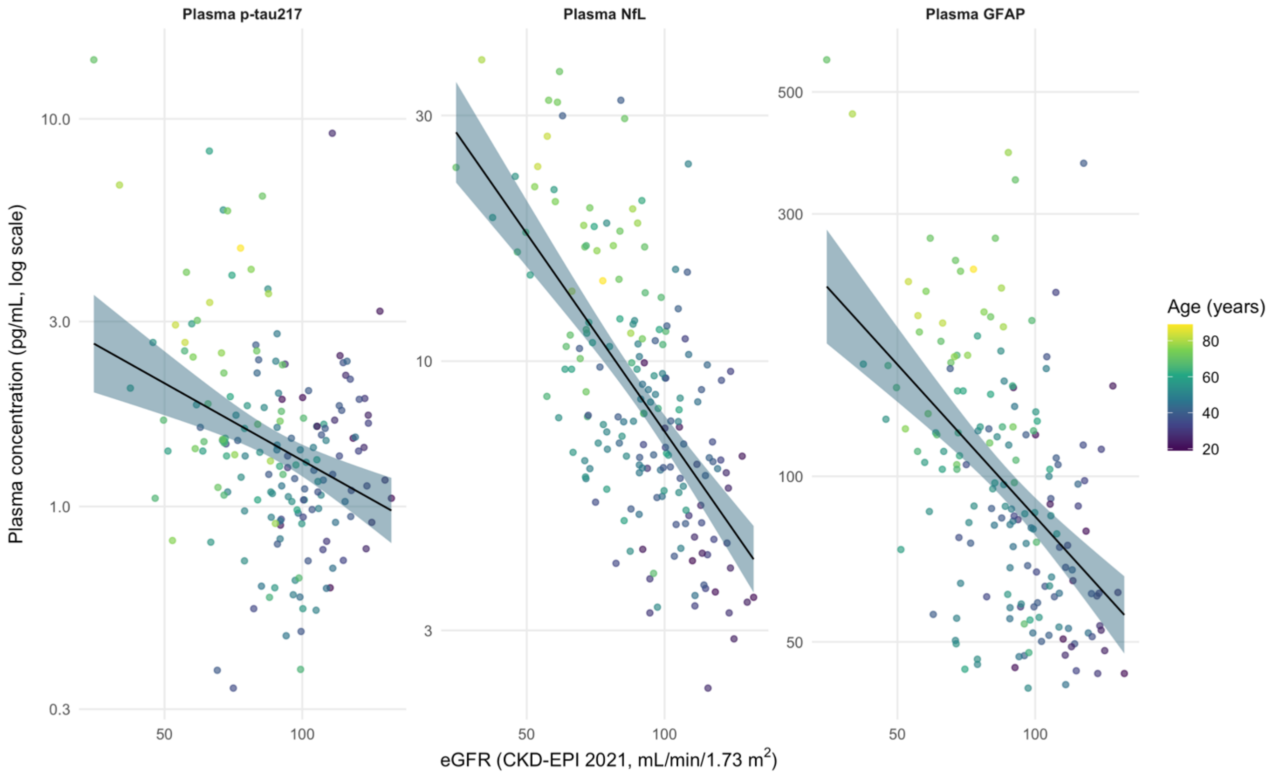


**eFigure 1.** Plasma p-tau217, NfL, and GFAP concentrations against eGFR (CKD-EPI 2021), coloured by age at blood draw. Fitted lines are unadjusted linear models on log-transformed concentrations, with 95% confidence bands. Concentrations are plotted on a logarithmic scale.
